# Long term follow-up in anti-contactin-1 autoimmune nodopathy

**DOI:** 10.1101/2024.06.25.24309231

**Authors:** Marta Caballero-Ávila, Lorena Martín-Aguilar, Elba Pascual-Goñi, Milou R. Michael, Marleen J.A. Koel-Simmelink, Romana Höftberger, Julia Wanschitz, Alicia Alonso-Jiménez, Thais Armangué, Adája Elisabeth Baars, Álvaro Carbayo, Barbara Castek, Roger Collet-Vidiella, Jonathan De Winter, Maria Angeles del Real, Emilien Delmont, Luca Diamanti, Pietro Emiliano Doneddu, Fu Liong Hiew, Eduard Gallardo, Amaia Gonzalez, Susanne Grinzinger, Alejandro Horga, Stephan Iglseder, Bart C. Jacobs, Amaia Jauregui, Joep Killestein, Elisabeth Lindeck Pozza, Laura Martínez-Martínez, Eduardo Nobile-Orazio, Nicolau Ortiz, Helena Pérez-Pérez, Kai-Nicolas Poppert, Paolo Ripellino, Jose Carlos Roche, Franscisco Javier Rodriguez de Rivera, Kevin Rostasy, Davide Sparasci, Clara Tejada-Illa, Charlotte C.E. Teunissen, Elisa Vegezzi, Tomàs Xuclà-Ferrarons, Fabian Zach, Luuk Wieske, Filip Eftimov, Cinta Lleixà, Luis Querol

## Abstract

**Objective:** To analyze long-term clinical and biomarker features of anti-contactin-1 (CNTN1) autoimmune nodopathy (AN).

**Methods:** Patients with anti-CNTN1+ AN detected in our laboratory from which clinical information was available were included. Clinical features and treatment response were retrospectively collected. Autoantibody, serum neurofilament light (sNfL) and serum CNTN1 levels (sCNTN1) were analyzed at baseline and follow-up.

**Results:** Thirty-one patients were included. Patients presented with progressive motor-sensory neuropathy (76.7%) with proximal (74.2%) and distal involvement (87.1%), ataxia (71.4%) and severe disability (median INCAT at nadir of 8)). Eleven patients (35%) showed kidney involvement. Most patients (97%) received IVIg but only one achieved remission with IVIg. Twenty-two patients (71%) received corticosteroids, and three of them (14%) did not need further treatments. Rituximab was effective in 21/22 patients (95.5%), with most of them (72%) receiving a single course. Four patients (12.9%) relapsed after a median follow-up of 25 months after effective treatment [12-48]. Anti-CNTN1 titers correlated with clinical scales at sampling and were negative after treatment in all patients but one (20/21). sNfL levels were significantly higher and sCNTN1 significantly lower in anti-CNTN1+ patients than in healthy controls (sNfL: 135.9 pg/mL vs 7.48 pg/mL, sCNTN1: 25.03 pg/mL vs 22186 pg/mL, p< 0.0001). Both sNfL and sCNTN1 returned to normal levels after successful treatment.

**Interpretation:** Patients with anti-CNTN1+ AN have a characteristic clinical profile. Clinical and immunological relapses are infrequent after successful treatment, suggesting that continuous treatment is unnecessary. Anti-CNTN1 antibodies, sNfL and aCNTN1 levels are useful to monitor disease status and treatment efficacy in these patients.

## 1. Introduction

Autoimmune nodopathies (AN) are a group of immune-mediated neuropathies associated with antibodies against cell adhesion molecules of the node of Ranvier (1,2). These antibodies target proteins such as contactin 1 (CNTN1) (3), contactin-associated protein 1 (Caspr1) (4,5), neurofascin 155 (NF115)(6,7) or pan-neurofascin (pan-NF) (8,9). They account for 5-10% of patients fulfilling diagnostic criteria of chronic inflammatory demyelinating polyradiculoneuropathy (CIDP)(10) However, they have recently been classified as a different diagnostic category in the recent update of the European Academy of Neurology/ Peripheral Nerve Society CIDP diagnostic guidelines(11) due to their specific clinical, pathological, and response to treatment profiles, that differ from those of CIDP.

CNTN1 is an axonal protein present in the paranodal region, where it forms a heterodimer with Caspr1 that binds to NF155 in the Schwann cell side of the paranode. This complex forms septate-like axoglial junctions, also known as transverse bands, that connect the paranodal loops of the Schwann cells to the axon (12,13). Autoantibodies against CNTN1 have been described in patients with rapidly progressive sensory-motor neuropathy with predominant distal weakness and sensory ataxia; and poor response to intravenous immunoglobulins (IVIg)(3,14,15). These antibodies have also been associated with membranous glomerulonephritis (MGN), that can appear concomitant to the neuropathy or isolated(16,17). Small case-series suggest that rituximab is effective in anti-CNTN1+ AN patients (18), but data on long-term follow-up and biomarker dynamics that help disease monitoring are lacking.

In this study we describe the clinical phenotype, long-term follow-up and response to treatment of the largest cohort of anti-CNTN1+ AN patients so far. We also report data on autoantibody, serum neurofilament light (sNfL) and serum CNTN1 (sCNTN1) levels that support the use of these biomarkers in the follow-up of anti-CNTN1+ AN patients.

## 2. Methods

### 2.1. Patients and samples

We included all patients with anti-CNTN1 antibodies identified during routine clinical testing of nodal/paranodal antibodies from which clinical information wasavailable. The samples were obtained between April 2002 and February 2024. These patients were selected for further characterization between May 2023 and September 2023. Demographic and clinical data were collected in a coded database. This study was conducted according to a protocol approved by the Ethics Committee of the Hospital de la Santa Creu i Sant Pau (code IIBSP-NAI-2022-88). All patients gave written informed consent to participate in the study.

Clinical and demographic data were retrospectively collected by chart review by treating neurologists. Information about ancillary tests (lumbar puncture, nerve biopsy) was collected when available. Disability scores, including the modified Rankin Scale (mRS, 0-6)(19), the Inflammatory Neuropathy Cause and Treatment (INCAT, 0-10)(20), and the inflammatory Rasch-built Overall Disability Scale (iRODS, 0-48) were collected at nadir and at follow-up when available (at 1 month, 3 months, 6 months and last follow-up). Response to therapy was defined as a good response, partial response, or no response as classified by their primary neurologists after chart review of the neurologic examination. Relapse was defined as clinical worsening after effective treatment.

Serum samples were obtained at diverse time points during routine autoantibody testing and stored at −80°C until needed.

### 2.2. Anti-CNTN1 antibody detection and titration

Antibodies against CNTN1 were analyzed by both fixed and live cell-based assay (CBA). A mammalian expression vector encoding full-length human CNTN1 cDNA (EX-A1153-M02, Genecopoeia, Maryland, USA) was transfected into HEK293 cells using lipofectamine 2000 (Invitrogen, CA, USA).

In fixed CBA, cells were then fixed with paraformaldehyde 4% and blocked with rabbit serum 1/40 in PBS. Double immunocytochemistry (ICC) was performed using 1/100 diluted sera or 1/10 diluted CSF, followed by anti-CNTN1 antibody at 1/1000 (AF904, R&D Systems, Minnesota, USA), and then secondary antibodies at 1/500 (rabbit anti-goat AF488 and rabbit anti-human AF594). Finally, slides were mounted with Fluoromount (Sigma, MO, USA) and examined by two independent observers. Images were obtained with an Olympus BX51 Fluorescence Microscope (Olympus Corporation, Tokyo, Japan).

In live CBA, sera diluted in 1/100 in cell culture medium were incubated with CNTN1-transfected HEK293 cells for one hour at 37°C. Then, cells were fixed with paraformaldehyde 4% and blocked with rabbit serum 1/40 in PBS. Primary and secondary antibodies were then incubated as in fixed CBA.

ELISA was used as a confirmatory technique and for titration and isotype identification. Maxisorb 96-well ELISA plates (Thermo Fisher Scientific, NUNC, Denmark) were coated overnight with 1 μg/ml human recombinant CNTN1 protein (Sino Biological Inc., Georgia, USA). Wells were blocked with 5% non-fat milk in PBS 0.1% Tween20 for 1h, incubated with sera diluted 1/100 for 1h, and then incubated with peroxidase conjugated rabbit anti-human IgG secondary antibody or IgG subtypes (Invitrogen) for 1h at room temperature. ELISA was developed with tetramethylbenzidine solution (BioLegend, CA, USA), and the reaction was stopped with 25% sulfuric acid. Optical density (OD) was measured at 450nm in a Multiscan ELISA reader. Samples were considered positive by ELISA when they had a ΔOD higher than the average healthy control ΔOD plus two standard deviations. To calculate the antibody titres, the ELISA was performed with different serum concentrations (range 1/100-1/24300).

### 2.3. sNfL levels detection

sNfL levels were measured in all available anti-CNTN1+ AN samples at onset and follow up, and compared with 60 aged-matched healthy controls (HC) and 111 Guillain-Barré syndrome (GBS) using the Simoa NF-light kit in the SR-X Immunoassay Simoa analyzer (Quanterix Corp, Boston, MA), as previously described (21). The samples were analyzed in duplicates following the manufacter’s instructions and standard procedures. All sNfL values were within the linear ranges of the assay. The intraassay and interassay coeficients of variation (CV) at intermediate level were 12.6% and 5.2%. sNfL percentiles and z-scores were calculated using the sNfL Reference App for both adults and children population (22,23). A z-score cutoff of 1.5 was considered relevant based on data published in multiple sclerosis (22).

### 2.4. sCNTN1 levels

sCNTN1 levels were measured in all available anti-CNTN1+ AN samples at onset and follow up, and compared with 74 healthy controls (HCs). sCNTN1 levels were measuredat the Neurochemistry Laboratory at Amsterdam UMC using the Human Magnetic Luminex Assay (LXSAHM; R&D Systems, Minneapolis, MN) on the Bio-Plex 200 system (Bio-Rad Laboratories, Veenendaal, The Netherlands) according to the manufacturer’s instructions as previously described (24). The intra-assay CV was 1.9% and none of the measurements had an intra-assay CV > 15%.

### 2.5. Statistical analysis

Descriptive statistics are shown as mean (±SD) or median [inter-quartile range] in continuous variables and as frequencies (percentage) in categorical variables. Differences in sNfL, sCNTN1 levels and clinical scales between groups were analyzed with the t-test or the Mann–Whitney U test when appropriate. Correlations between anti-CNTN1 antibody titers or sNfL and clinical scales were assessed using the Spearman coefficientThe Kruskall Wallis test was used to compare sNfL levels across groups based on anti-CNTN1 antibody titers. Statistical significance for all analyses was set at 0.05 (2-sided). All statistical analysis and graphs were done with SPSS version 26 (IBM Corp) and GraphPad Prism v9.

## 3. Results

### 3.1. Anti-CNTN1 antibody screening

Thirty-two patients positive for anti-CNTN1 antibodies from 2002 to 2023 were identified. However, positivity could not be confirmed in one patient, resulting in thirty-one anti-CNTN1+ patients included in the study. All serum samples were positive in fixed and live CNTN1 CBA, except one that was only positive in live CBA (Supplementary Figure 1). In 10 patients, only the initial sample was available; samples at different timepoints were available in 21 (11 patients with 2 samples, 3 patients with 3, 3 patients with 4, 1 patient with 6, 1 patient with 9). Cerebrospinal fluid (CSF) was available in 3 patients.

### 3.2. Baseline clinical features

Mean age at disease onset was 50.3 years and 24 patients (77.4%) were male. Initial diagnosis was GBS in 16 (51.6%) patients, CIDP in 12 (38.7%) and others in 3 (sensory neuropathy, lumbosacral radiculopathy and diabetic neuropathy). Three (9.35%) patients were children. The most common form of presentation was sensory-motor (23/30, 76.7%), with 5 patients that present with a sensory-ataxic form (16.7%) and two with a pure motor form (6.7%). Most patients had symmetrical (23/29, 79.3%) weakness. Distal weakness was present in 27 patients (87.1%; upper limbs: 24/31, 77.4%; lower limbs: 23/31 74.2%), and 23 patients (74.2%) had proximal weakness (upper limbs: 15/31, 48.4%; lower limbs: 23/31 74.2%), Most patients had sensory deficit in the lower limbs (28/31, 93.5%) and 23 in the upper limbs (74.2%). Ataxia was frequent (20/28, 71.4%), being severe (walking aids needed) in 13 patients (46.4%); seven patients (24.1%) had tremor. Facial weakness was the most common cranial involvement (8/31, 25.8%), and respiratory involvement was rare (2/31, 6.5%).

Lumbar puncture was performed in 30 patients and revealed a median protein level in CSF of 2.4 g/L [1.4-5.8]. Nerve biopsy (sural) was obtained in five patients (3 axonal neuropathy, 1 demyelinating neuropathy, 1 normal).

Eleven patients (35.5%) had a renal disease. Five patients (16.1%) were diagnosed with membranous glomerulonephritis (MGN) during or after neuropathy onset; one of them had proteinuria before neuropathy onset. Three patients had nephrotic range proteinuria (but a kidney biopsy was not done). Three additional patients had a previous kidney involvement of unknown cause (2 chronic kidney disease and 1 post-renal acute failure).

Further information on clinical features is detailed in Table 1.

**Table 1.**
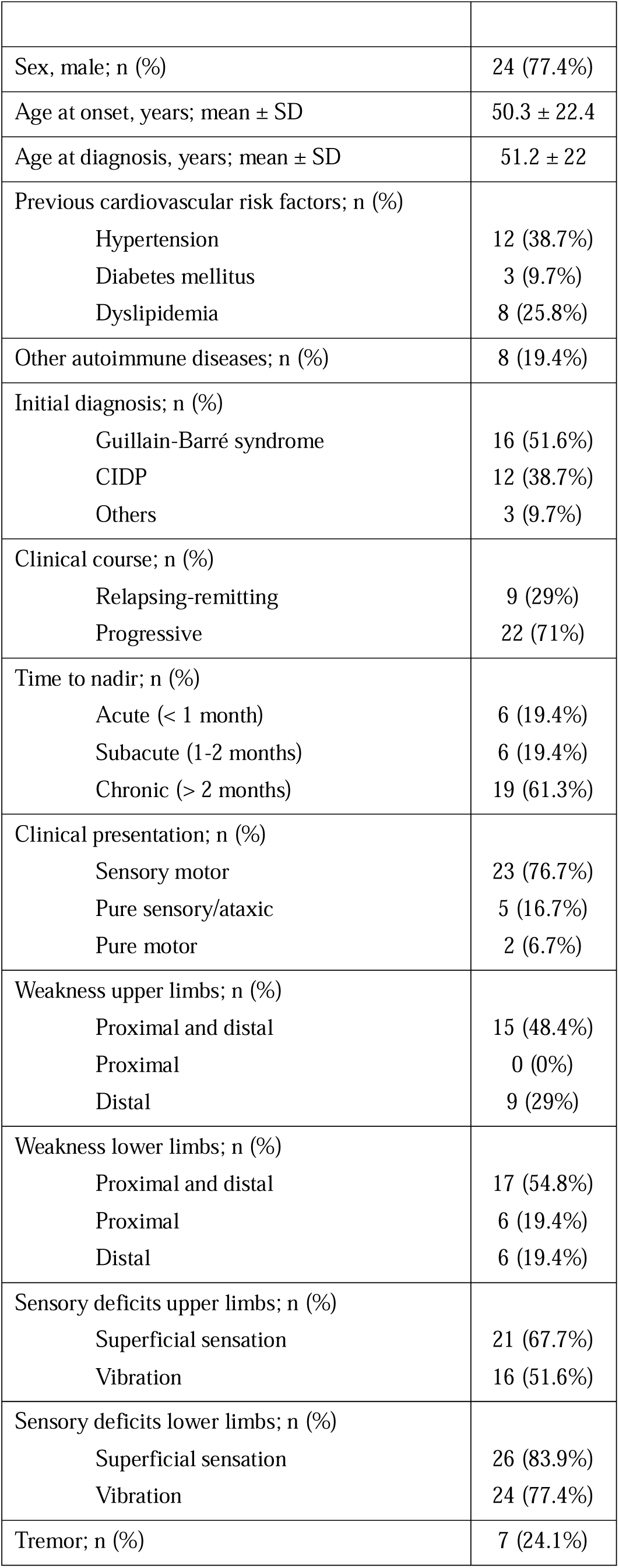
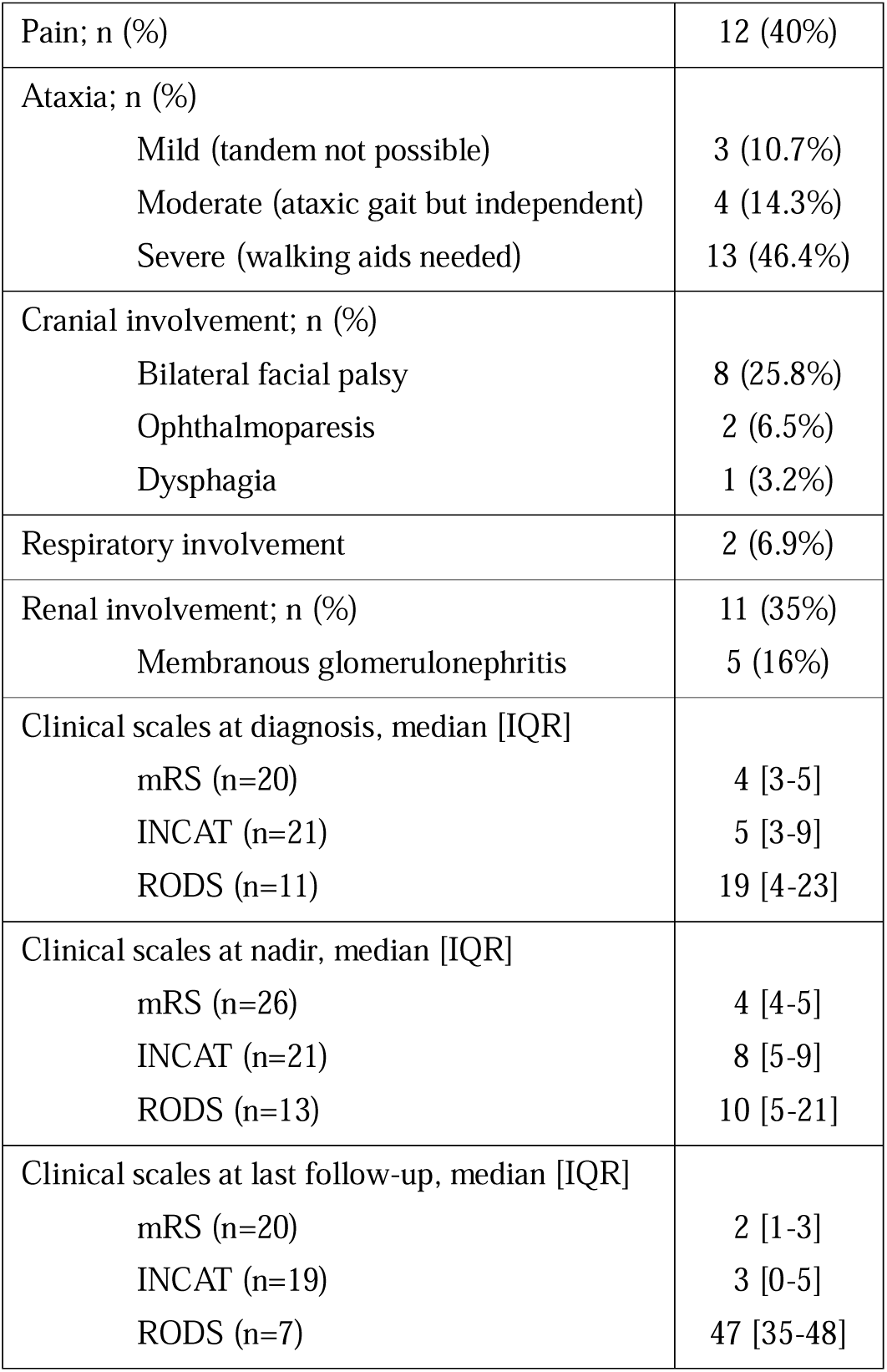
Baseline demographic and clinical data of anti-CNTN1 AN patients (n=31)

|  |  |
| --- | --- |
| Sex, male; n (%) | 24 (77.4%) |
| Age at onset, years; mean ± SD | 50.3 ± 22.4 |
| Age at diagnosis, years; mean ± SD | 51.2 ± 22 |
| Previous cardiovascular risk factors; n (%) |  |
| Hypertension | 12 (38.7%) |
| Diabetes mellitus | 3 (9.7%) |
| Dyslipidemia | 8 (25.8%) |
| Other autoimmune diseases; n (%) | 8 (19.4%) |
| Initial diagnosis; n (%) |  |
| Guillain-Barré syndrome | 16 (51.6%) |
| CIDP | 12 (38.7%) |
| Others | 3 (9.7%) |
| Clinical course; n (%) |  |
| Relapsing-remitting | 9 (29%) |
| Progressive | 22 (71%) |
| Time to nadir; n (%) |  |
| Acute (< 1 month) | 6 (19.4%) |
| Subacute (1-2 months) | 6 (19.4%) |
| Chronic (> 2 months) | 19 (61.3%) |
| Clinical presentation; n (%) |  |
| Sensory motor | 23 (76.7%) |
| Pure sensory/ataxic | 5 (16.7%) |
| Pure motor | 2 (6.7%) |
| Weakness upper limbs; n (%) |  |
| Proximal and distal | 15 (48.4%) |
| Proximal | 0 (0%) |
| Distal | 9 (29%) |
| Weakness lower limbs; n (%) |  |
| Proximal and distal | 17 (54.8%) |
| Proximal | 6 (19.4%) |
| Distal | 6 (19.4%) |
| Sensory deficits upper limbs; n (%) |  |
| Superficial sensation | 21 (67.7%) |
| Vibration | 16 (51.6%) |
| Sensory deficits lower limbs; n (%) |  |
| Superficial sensation | 26 (83.9%) |
| Vibration | 24 (77.4%) |
| Tremor; n (%) | 7 (24.1%) |
| Pain; n (%) | 12 (40%) |
| Ataxia; n (%) |  |
| Mild (tandem not possible) | 3 (10.7%) |
| Moderate (ataxic gait but independent) | 4 (14.3%) |
| Severe (walking aids needed) | 13 (46.4%) |
| Cranial involvement; n (%) |  |
| Bilateral facial palsy | 8 (25.8%) |
| Ophthalmoparesis | 2 (6.5%) |
| Dysphagia | 1 (3.2%) |
| Respiratory involvement | 2 (6.9%) |
| Renal involvement; n (%) | 11 (35%) |
| Membranous glomerulonephritis | 5 (16%) |
| Clinical scales at diagnosis, median [IQR] |  |
| mRS (n=20) | 4 [3-5] |
| INCAT (n=21) | 5 [3-9] |
| RODS (n=11) | 19 [4-23] |
| Clinical scales at nadir, median [IQR] |  |
| mRS (n=26) | 4 [4-5] |
| INCAT (n=21) | 8 [5-9] |
| RODS (n=13) | 10 [5-21] |
| Clinical scales at last follow-up, median [IQR] |  |
| mRS (n=20) | 2 [1-3] |
| INCAT (n=19) | 3 [0-5] |
| RODS (n=7) | 47 [35-48] |

### 3.3. Clinical response to treatment and follow-up

Patients received a median of 3 [2-4] treatments during follow-up. Data on patient’s treatment and response are summarized in Table 2. All patients except one received IVIg (97%), 22 patients received corticosteroids (71.0%), and 15 patients (48.4%) were treated with plasma exchange (PLEX), with a median 5 sessions [4-5]. Twenty-two patients (71.0%) were treated with rituximab. Cyclophosphamide was used in three patients (9.8%), cyclosporine in 1 patient (3.2%) and azathioprine in 1 patient (3.2%).

**Table 2.** Treatment and clinical response of anti-CNTN1 AN patients.

|  | **Number patients (n,%)** | **Response, (n,%)** | **Dose/Protocol, (n,%)** |
| --- | --- | --- | --- |
| **IVIg** | 30 (97%) | Yes: 6 , 20%<br>Partial: 13, 43.3%<br>No: 11, 36.7% |  |
| **PLEX** | 15 (48.4%) | Yes: 5, 33.3%<br>Partial: 8, 53.3%<br>No: 2, 13.3% | Num sessions, median [IQR]: 5 [4-5] |
| **Corticosteroids** | 22 (71%) | Yes: 8, 36.4%<br>Partial: 6, 27.3%<br>No: 8, 36.4% | MP pulses: 10, 45.4%<br>mg/kg: 10, 45.4%<br>Others: 3, 13.6% |
| **Rituximab** | 22 (71%) | Yes: 16, 72.7%<br>Partial: 5, 22.7%<br>No: 0 | 1+1: 13, 59%<br>4: 2, 9%<br>4+2: 1, 4.5%<br>Others: 6, 27.3% |
| **Last effective treatment** |  |  |  |
| IVIg | 1, 3.2% |  |  |
| IVIg+ corticosteroids | 2, 6.5% |  |  |
| Corticosteroids | 3, 9.7% |  |  |
| Rituximab | 21, 67.7% |  |  |
| No effective treatment | 3, 9.7% |  |  |

Six patients had good response to IVIg (20%) and thirteen (43.3%) partial response. Only one patient with good response to IVIG (3.3%) did not need additional therapies. All other patients required treatment with corticosteroids and/or rituximab due to neuropathy worsening. Response to corticosteroids was good in 8 patients (36.7%) and partial in six patients (27.2%). Three (13.6%) of the patients that responded to corticosteroids did not need additional therapies. Two (6.5%) received IVIg combined with corticosteroids to achieve neuropathy remission and it was not possible to assign the response to any of them. Three patients needed rituximab as a second line therapy despite showing good initial response to corticosteroids. Most common corticosteroid regimens were oral prednisone 1mg/kg/day (10/22, 45.4%) and oral or intravenous methylprednisolone pulses (10/22, 45.4%).

Rituximab was effective in 16 patients (16/22, 72.7%), and partially effective in 5 (5/22, 22.7%). One patient’s response to rituximab is pending evaluation due to recent administration. However, antibody titers have become negative and sNfL have decreased significantly in this patient. The most common infusion protocol was two doses of 1000 mg (13/22, 59.1%); Sixteen patients received a single course of treatment (two patients received 2courses, and 3 patients every 6 months). Nine patients were not treated with rituximab. Reasons of no treating with rituximab were: 6 patients with good response to corticosteroids and/or IVIg, two patients died before starting rituximab, and reason in one is unknown.

Clinical scales at diagnosis, nadir and last follow-up are summarized in Table 1. Worst scores on clinical scales (mRS, INCAT and RODS) were detected at nadir (Table 1) with significant improvement during follow-up (mRS, INCAT and RODS at nadir vs last follow-up, p<0.0001) (Figure 1). The median follow-up time was 35 months [18-76] from onset and 25 months [12-48] from effective treatment (Figure 2). Four patients (12.9%) experienced a clinical and immunological relapse after effective treatment: one patient treated with corticosteroids and three with rituximab. The first one was retreated with corticosteroids and achieved remission. In rituximab-treated patients, relapse appeared after a median time from infusion of 18 months [16-55]. Two of them were retreated with rituximab (one with good response, other pending evaluation), and the other one with rituximab and cyclophosphamide with no response. Finally, another patient worsened two months after starting rituximab (and needed adding corticosteroids) but was not considered a relapse because it had been a short time since the start of rituximab. Four (12,9%) patients died during follow-up: two because of the neuropathy, one of pneumonia and another of a disseminated neoplasm.

**Figure 1.**
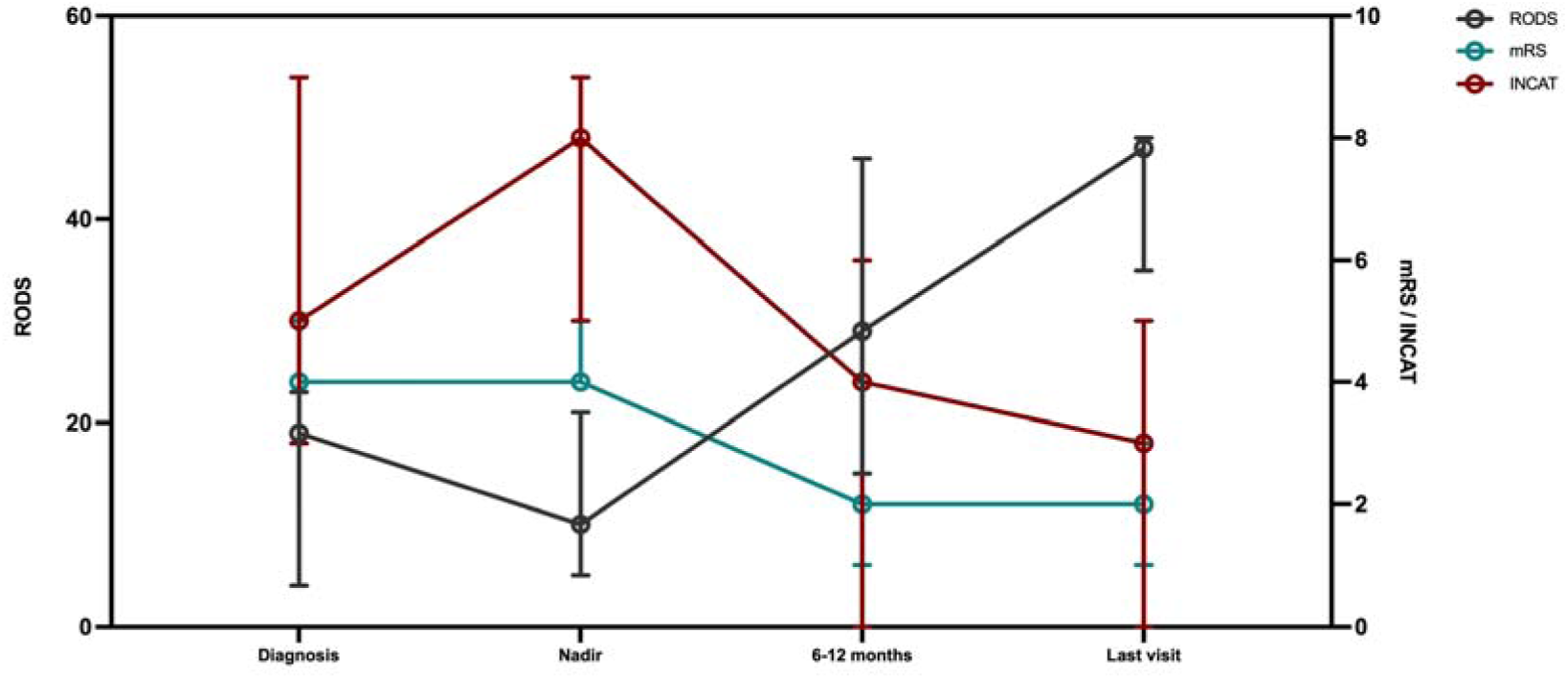
Clinical scales of anti-CNTN1+ AN patients at onset and follow-up. Patients in which clinical scales were available at diagnosis, nadir, 6-12 months and last visit were: 26, 26, 17, 20 for mRS; 21, 21, 15, 19 for INCAT; and 11, 13, 9, 7 for RODS. The circle represents the median value and the wiskers indicate the interquartile range.

**Figure 2.**
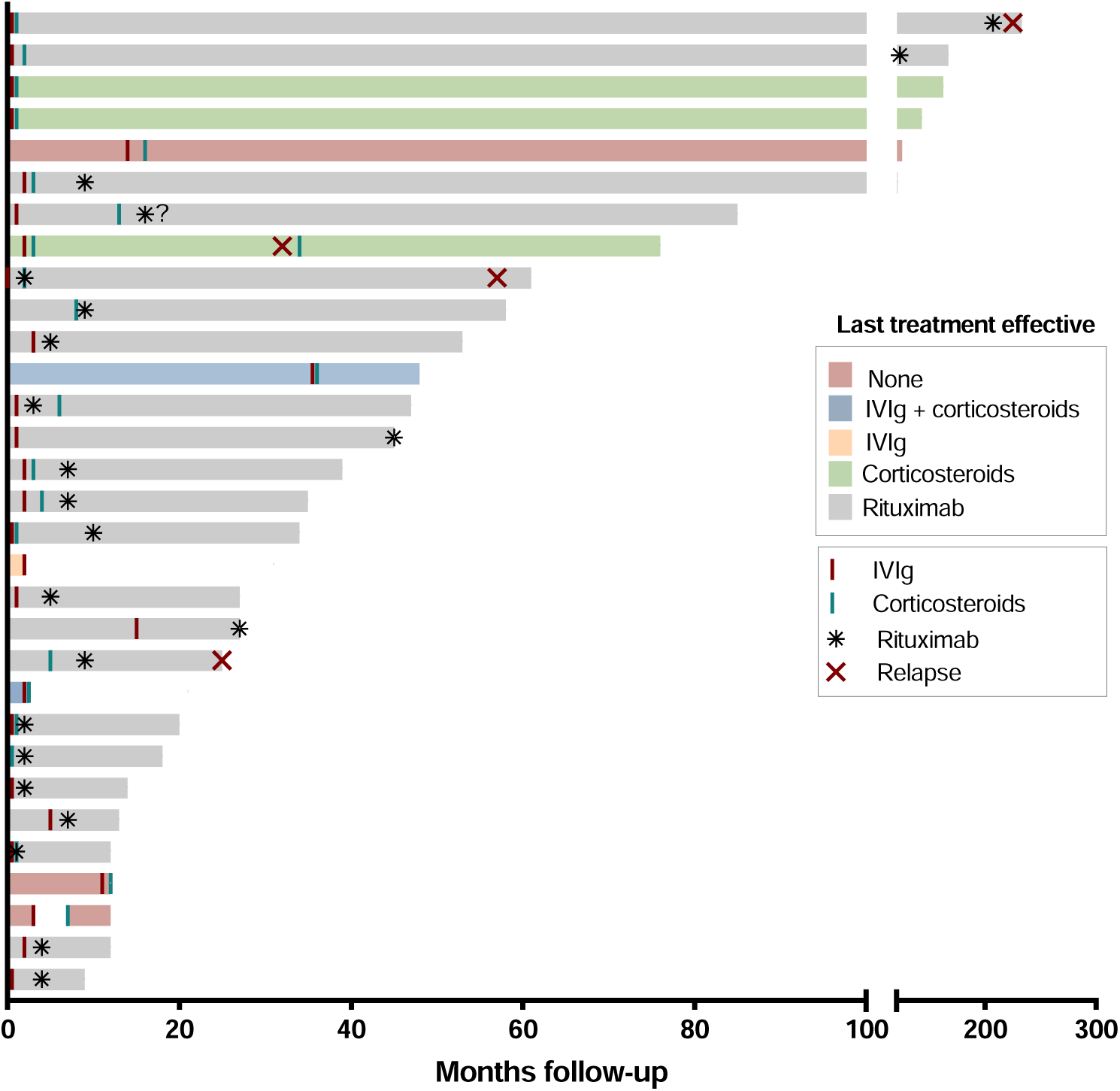
Follow-up of all anti-CNTN1+ patients (n=31). Patients are represented individually from onset to last follow-up. Treatment with IV immunoglobulins (IVIg), corticosteroids, rituximab and relapses are indicated as expressed in the legend. Bar color indicate last treatment effective. Exact date of infusion of rituximab in one patient is missing (expressed as a question mark).

### 3.4 Immunological characteristics

Anti-CNTN1 CBA positivity was confirmed in all patients by ELISA. Titers of anti-CNTN1 antibodies at first sample ranged from 1/900 to >1/24300. Autoantibodies were predominantly of the IgG4 subclass in all patients but one (IgG1 only). All patients (n=21) were negative for anti-CNTN1 antibodies after treatment, except for one patient with high anti-CNTN1 titers (>1/24300) tested 6 months after rituximab that remained positive at significantly lower titers (1/900). All patients that presented with a clinical relapse after rituximab were positive at the moment of relapse for anti-CNTN1 antibodies. Finally, we were able to study the presence of anti-CNTN1 antibodies in CSF from three patients by fixed CBA and ELISA, with only one patient testing positive.

### 3.5. sNfL

sNfL levels of anti-CNTN1+ AN patients were analyzed in samples available at onset (n=28) and at last follow-up (n=21); in 111 GBS patients and 60 HCs (Figure 3A). Additionally, sNfL were also analyzed in other moments of the disease in 10 patients to analyze the kinetics (n=19). sNfL levels were significantly higher at onset compared with last follow-up (135.9 pg/mL vs 9.59 pg/mL, p<0.0001) and with HC (7.48 pg/mL, p<0.0001). sNfL at onset were also significantly higher compared with GBS patients (48.04 pg/mL, p=0.004). No differences were found between sNfL in anti-CNTN1+ AN patients at last follow-up and healthy controls (p=0.6). Regarding z-score, all patients before treatment had a z-score above 1.5, with normalization after treatment (z-score <1.5) in 17/21 patients (81%) (Figure 3B). Four patients remained with a z-score >1.5 after treatment, however, three of them started in significantly higher z-scores (1.5-fold), and in one of them onset sample is missing.

**Figure 3.**
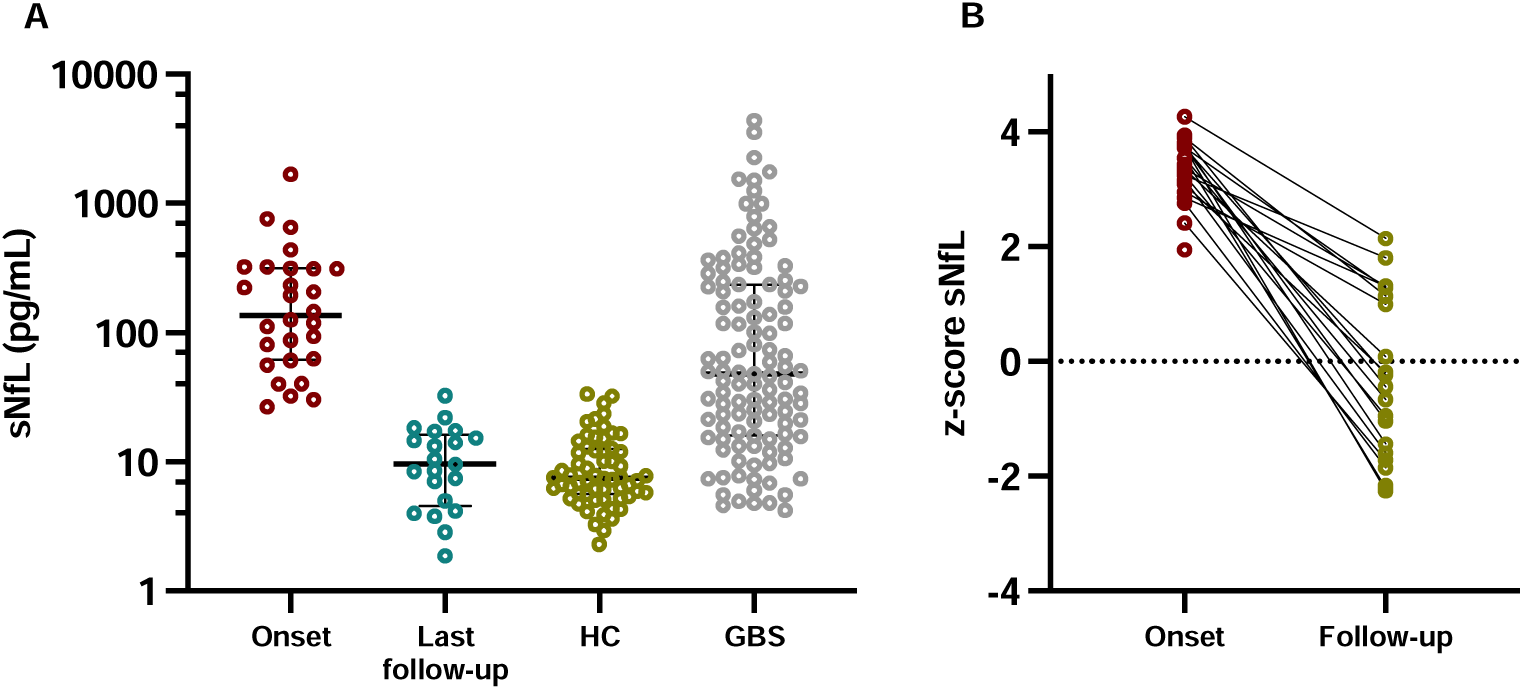
Patients with anti-CNTN1+ AN had significantly higher sNfL at onset compared to last follow-up, HC and GBS patients. Figure 2A is showing raw values while figure 2B is showing z-scores. In Figure 2A, the line in the center represents the median value and the wiskers indicate the interquartile range. HC= healthy controls; GBS= Guillain-Barré Syndrome; sNfL= serum neurofilament light chain.

### 3.6. sCNTN1 levels

sCNTN1 levels were analyzed in 21 anti-CNTN1+ AN patients (13 patients with samples at onset and follow-up, 3 patients only at onset and 5 patients only at last follow-up), and in 74 healthy controls. Age was not taken into consideration since sCNTN1 has not been associated with age(24). Median sCNTN1 at onset was 25.03 pg/mL [7.6-180.6] and was significantly lower compared to samples at last follow up (median 21801 pg/mL [2930-36779], p=0.0003), and to healthy controls (median 22186 pg/mL [19729-24735], p<0.0001) (Figure 4). No differences were found between samples at last follow-up and healthy controls (p=0.85), suggesting that patients after treatment achieve normal levels of sCNTN1. Five follow-up samples had low levels of sCNTN1 (<10000 pg/mL). These follow-up samples were collected a median of 7 months (2–8) after onset, a significantly earlier follow-up sample than the patients with sCNTN1 increase (median 27.5 months [10.5-74.6], p=0.01).

**Figure 4.**
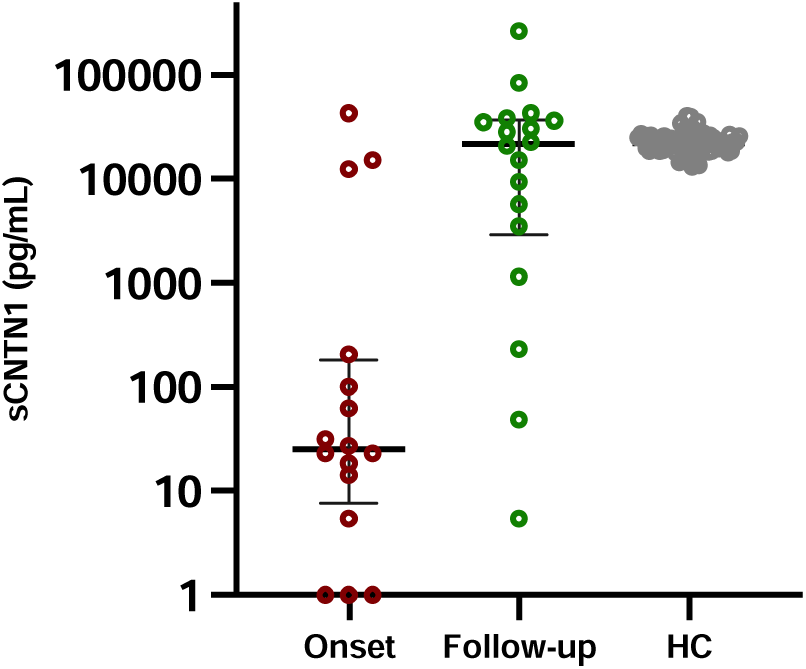
Serum CNTN1 levels. Patients with anti-CNTN1+ AN at onset had significantly lower levels of sCNTN1 levels compared to last follow-up and healthy controls (HC). The line in the center represents the median value and the wiskers indicate the interquartile range.

### 3.7. Relationship between biomarkersand clinical status

During follow-up all patients tested after treatment but one (1/21) were negative for anti-CNTN1 antibodies. sNfL decreased in all patients tested after treatment (n=21) achieving normal levels at last follow-up. Worst scores on clinical scales (mRS, INCAT and RODS) were detected at nadir (Table 1) with significant improvement during follow-up (mRS, INCAT and RODS at nadir vs last follow-up, p<0.0001). Median anti-CNTN1 titers, sNfL and sCNTN1 at different follow-up timepoints available are summarized in Figure 5.

**Figure 5.**
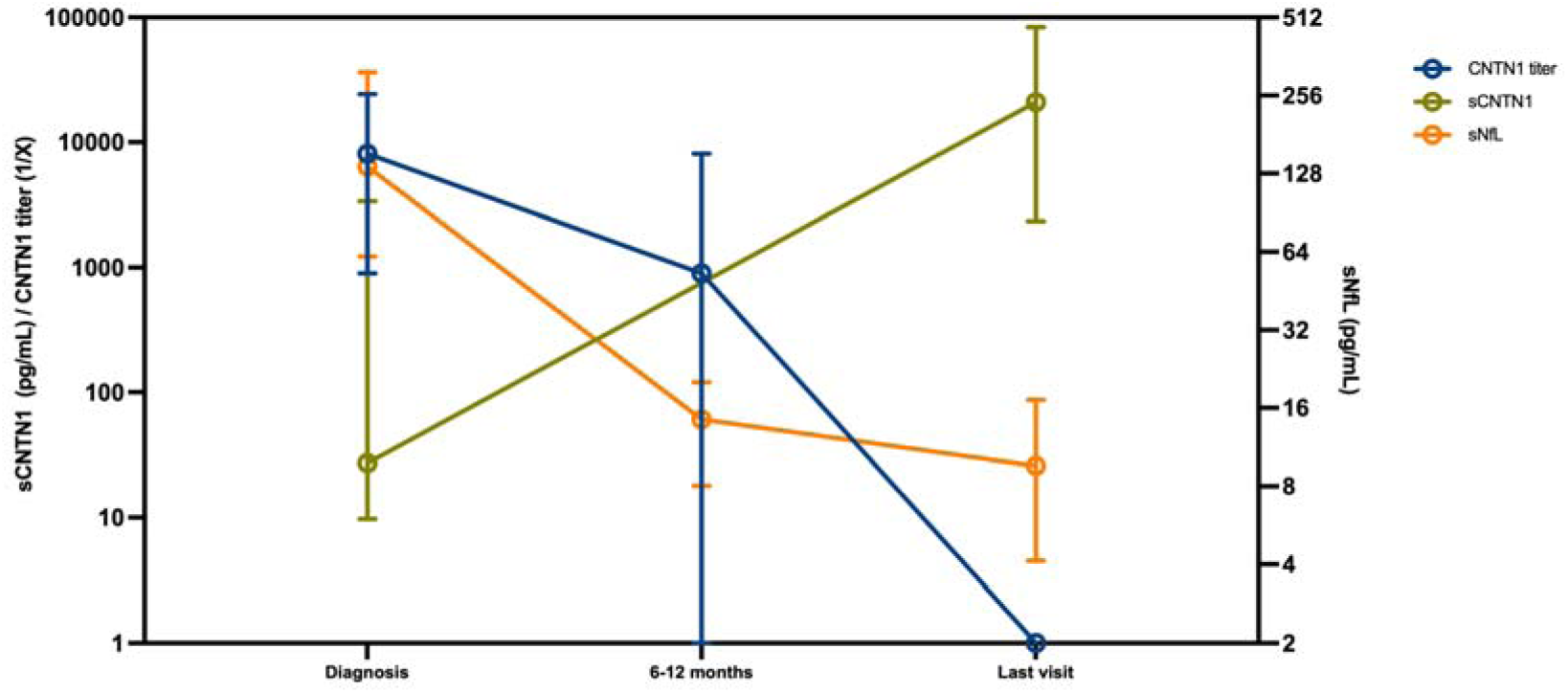
Biomarkers kinetics of anti-CNTN1+ AN patients at onset and follow-up. Samples available for sNfL were 28 at diagnosis, 14 at 6-12months, 15 at last visit; for CNTN1 titer were 27 at diagnosis, 14 at 6-12months, 15 at last visit; and form sCNTN1 were 17 at diagnosis and at last follow-up. The circle represents the median value and the wiskers indicate the interquartile range.

Absolute anti-CNTN1 titers at different points of the disease correlated with mRS (n=38, r= 0.61, p<0.0001), INCAT (n =43, r=0.58, p=0.0002) and sNfL (n=61, r=0.71, p<0.0001) at sampling, and negatively correlated with RODS also at sampling (n=20, r=-0.48, p=0.03) (Figure 6). sNfL also correlated with INCAT (n=38, r=0.64, p<0.0001) and negatively with RODS (n=22, r=-0.49, p=0.02) at sampling but not with mRS. sCNTN1 levels were correlated negatively with mRS at sampling (n=35, r=-0.42, p=0.01) and negatively with sNfL (n=30, r=- 0.0.66, p<0.0001). However, we did not find any correlation between antibody titers, sNfL or sCNTN1 levels at onset, with disability at last clinical evaluation.

**Figure 6.**
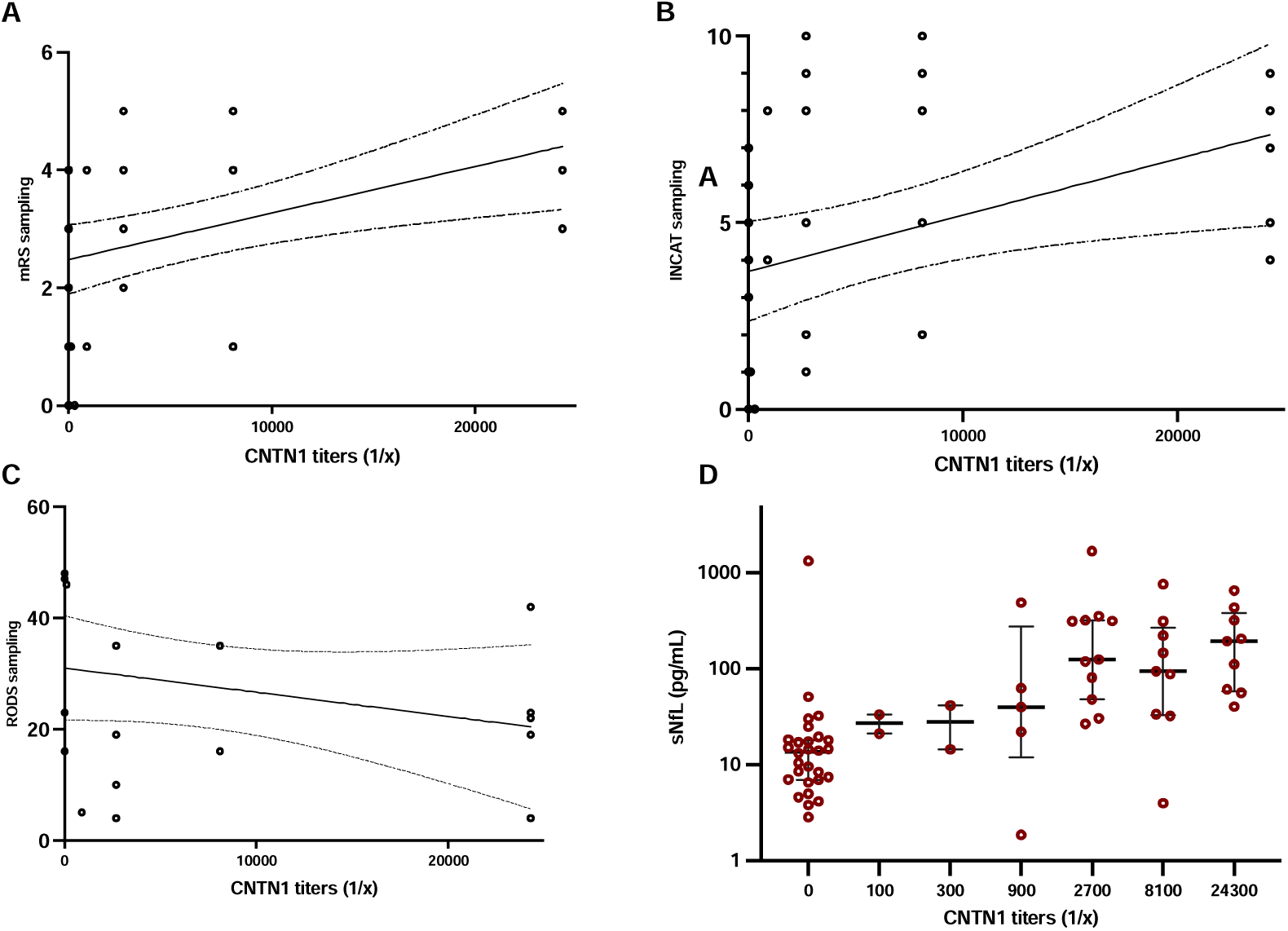
Clinical correlations of anti-CNTN1 titers. Antibody titers correlate with mRS (6A) and INCAT (6B) and sNfL (6D) at sampling, and negatively with RODS (6C).

## 4. Discussion

Our study describes the clinical and biomarker features of the largest anti-CNTN1+ AN cohort so far and supports that patients with anti-CNTN1 antibodies have a specific clinical phenotype that includes sensory-motor neuropathy, proximal and distal involvement and severe ataxia, combined with very high CSF protein levels and poor response to IVIg. Renal involvement is present in a third of the patients, highlighting the importance of studying the presence of nephropathy in these patients. Our data supports the utility of rituximab to achieve disease remission in this disorder, even after a single rituximab course, and reveals a low relapse rate, suggesting that continuous treatment might not be necessaryi most patients once remission is achieved. Finally, anti-CNTN1 antibody titers, sNfL levels and sCNTN1 levels are useful to monitor response to treatment and disease activity and may help in therapeutic decision making in the follow-up.

Anti-CNTN1+ AN is a very rare, but relevant, autoimmune neuropathy. As such, only small cohorts and case-series of patients with this conditionhave been published in the last years (14,15,25) and patients’ features and therapeutic regimens are based on limited information, particularly in the long-term. In the first description, anti-CNTN1+ AN patients were reported to be older than CIDP patients (3). Median age in our cohort was similar to that of other CIDP cohorts(26) and in fact, 10% of our patients were children. Information about pediatric AN is scarce, with a single cohort of children with inflammatory neuropathies that showed prevalence of antibodies to nodal/paranodal proteins can be similar to the adult population (27). Initially, it seemed that pediatric anti-CNTN1 could be less aggressive as in the first case published (28) rituximab was not needed to achieve remission. However, thehe two other pediatric patients included in our study received rituximab, suggesting that anti-CNTN1+ children may behave similar to adult cases. We have not found relevant associations with other medical conditions, including diabetes mellitus that had been previously associated to anti-CNTN1+ AN (29). Finally, very high CSF protein levels are found in these patients, so this could also be another feature to suspect the presence of the anti-CNTN1 antibodies.

About one third of the patients included in our study had renal disease, (half of them a MGN). The relationship between nephropathy and anti-CNTN1 antibodies was first described in isolated case reports or small case series (16,25,27,30–32). A recent study demonstrated the pathological association between MGN and neuropathy in patients with anti-CNTN1 antibodies(17). The study described the presence of CNTN1 protein in the renal tissue and reported anti-CNTN1+ patients with MGNin the absence of neuropathy. At least one third of the patients included in our study had clinical or/and biochemical signs of nephropathy. Frequency of kidney disease is lower than previously reported(32) but nevertheless highlights the importance of studying the presence of proteinuria and renal involvement in this group of patients.

We have also confirmed that IgG4 is the predominant autoantibody subtype in anti-CNTN1+ AN patients. As observed in other IgG4 related diseases (33), anti-CNTN1 AN patients respond less frequently and most often partially to IVIg, while most patients treated with rituximab display a good and long-lasting response. The role of corticosteroids in anti-CNTN1+ AN remains unclear. In other AN patients, response to corticosteroids is poor and limited to small group of patients(5,7,8). In our cohort, three patients achieved clinical and serological remission after corticosteroid treatment. A recent work presented two anti-CNTN1+ AN patients (34) with excellent response to corticosteroids. Because of that, we suggest that corticosteroids can be considered as a treatment option specially in patients that are not severely affected and are not rapidly progressing. Moreover, isolated case reports (35)or small series of patients(18) describing the beneficial effect of rituximab in anti-CNTN1+ AN patients have been previously published. A major contribution of our study is that most anti-CNTN1+ AN patients in which rituximab is administered only need one course to achieve long-lasting remission. Even if other therapies (corticosteroids, cyclophosphamide…) can also be useful, we suggest that rituximab should be considered as an early therapeutic option (or event as first-line treatment) in this disorder, since a single course could be enough to maximize disease control, minimizing side effects, treatment burdenand costs.

Anti-CNTN1 antibodies are pathogenic according to in vitro and in vivo studies (16,36–38). We hypothesized that antibody levels should correlate with clinical status. The relationship between clinical status and antibody titers is well known in some other IgG4 neurological diseases (7,8,39), but data on anti-CNTN1 antibodies do not exist. Importantly for monitoring purposes, our study supports that IgG4 anti-CNTN1 antibodies disappear in remission and re-appear again in relapses and, thus, antibody titers could be used to monitor disease activity and guide the need of further treatment.

Furthermore, our study also demonstrates that patients with anti-CNTN1+ AN have high levels of sNfL compared to healthy controls and even to GBS patients at onset. sNfL levels associate with disease severity and axonal variants, and have an independent prognostic value in GBS patients (21,40), suggesting that they could also be useful to detect axonal damage in autoimmune nodopathies. sNfL levels have been studied in anti-NF155 patients (7) and pan-NF (8), and have also been associated with disease activity. All patients included in our cohort showed elevated sNfL levels at onset when compared with normal levels stratified by age, and in most of them returned to normal levels upon successful treatment. This drop in sNfL was accompanied by clinical improvement in all cases, demonstrating that sNfL can also be used to monitor disease activity and need of treatment escalation. Finally, unlike in GBS, we have not found that sNfL at onset are associated with of long-term prognosis. This may be for two reasons, First: factors other than axonal damage, such as treatment choices, diagnostic delay and age of the patients, may also influence long-term disability; Second: the number of patients included in our cohort is not sufficient to demonstrate a prognosis value of sNfL, as it happens in GBS.

CNTN1 is a secreted protein that remains attached to the axonal membrane through glycosylphosphatidylinositol (GPI) anchors in the myelinated nerves, but CNTN1 can also be found in plasma in a soluble form (41). As CNTN1 is also expressed in myelinating fibers of the central nervous system, it was initially studied as a soluble biomarker in multiple sclerosis, demonstrating its prognostic value(42,43). Later, it was demonstrated that AN patients had markedly lower levels of sCNTN1 when compared with CIDP patients, being anti-CNTN1 AN patients the subgroup with the lowest levels of sCNTN1 (24). Although the logical hypothesis of this reduction was that anti-CNTN1 antibodies cleared sCNTN1, it was unclear because other AN like anti-NF155 had also low levels of sCNTN1, so this reduction could be related to other inherent factors of the disease. We now confirm that sCNTN1 levels are extremely low in CNTN1+ AN patients at onset, and return to normal levels after treatment, suggesting that the drop in the antibody titers plays a role on the reappearance of sCNTN1 in the anti-CNTN1 AN subgroup (not in the rest of AN). However, the return to normal levels of sCNTN1 seems slower than with sNfL so, although sCNTN1 might also be a good biomarker to monitor disease activity, its comparative advantage with sNfL is unclear.

The main limitation of our study is the small number of patients and its retrospective nature, including the retrospective analysis of treatment efficacy using chart review. More frequent follow-up visits using solid clinimetrics and sampling is necessary to better understand the rate of improvement in these severely disabled patients and the dynamics of anti-CNTN1 antibodies, sNfL and sCNTN1 to assess their usability as biomarkers of disease activity. Moreover, data on nerve conduction studies were not available for most patients, and a thorough neurophysiological phenotyping of these patients is still pending. Despite that, anti-CNTN1 AN is an extremely rare disorder and, with 31 patients, our cohort provides useful information on clinical, biomarker and treatment response features that will inform the neuromuscular community in how to treat these patients more efficiently.

In conclusion, our study supports that, first, anti-CNTN1 AN patients represent a distinct subset of autoimmune neuropathies with poor response to IVIg and excellent and long-lasting response to rituximab; second, that clinical and immunological relapses are rare in this disorder; and third that antibody titers, sNfL and sCNTN1 might be useful biomarkers to monitor disease activity.

## Data Availability

All data produced in the present study are available upon reasonable request to the authors

## Acknowledgments

This work was supported by by Fondo de Investigaciones Sanitarias (FIS), instituto de Carlos III, Spain, under grant PI22/00387. M. Caballero-Ávila was supported by a personal Rio Hortega grant CM21/00101. L. Martín-Aguilar was supported by a personal Juan Rodés grant JR21/00060. E. Pascual-Goñi was supported by the Benson Fellowship grant from the GBS-CIDP foundation. A. Carbayo was supported by a personal Rio Hortega grant CM21/00057. R Collet-Vidiella was supported by a personal Rio Hortega grant CM23/00002. Luis Querol was supported by a personal clinical intensification INT23/00066.

The authors acknowledge the Department of Medicine at the Universitat Auton oma de Barcelona. The authors also thank all our patients for their support and collaboration. Several authors of this publication are members of the European Reference Network for rare neuromuscular diseases (EURO-NMD).

---

**Supplementary Figure 1.**
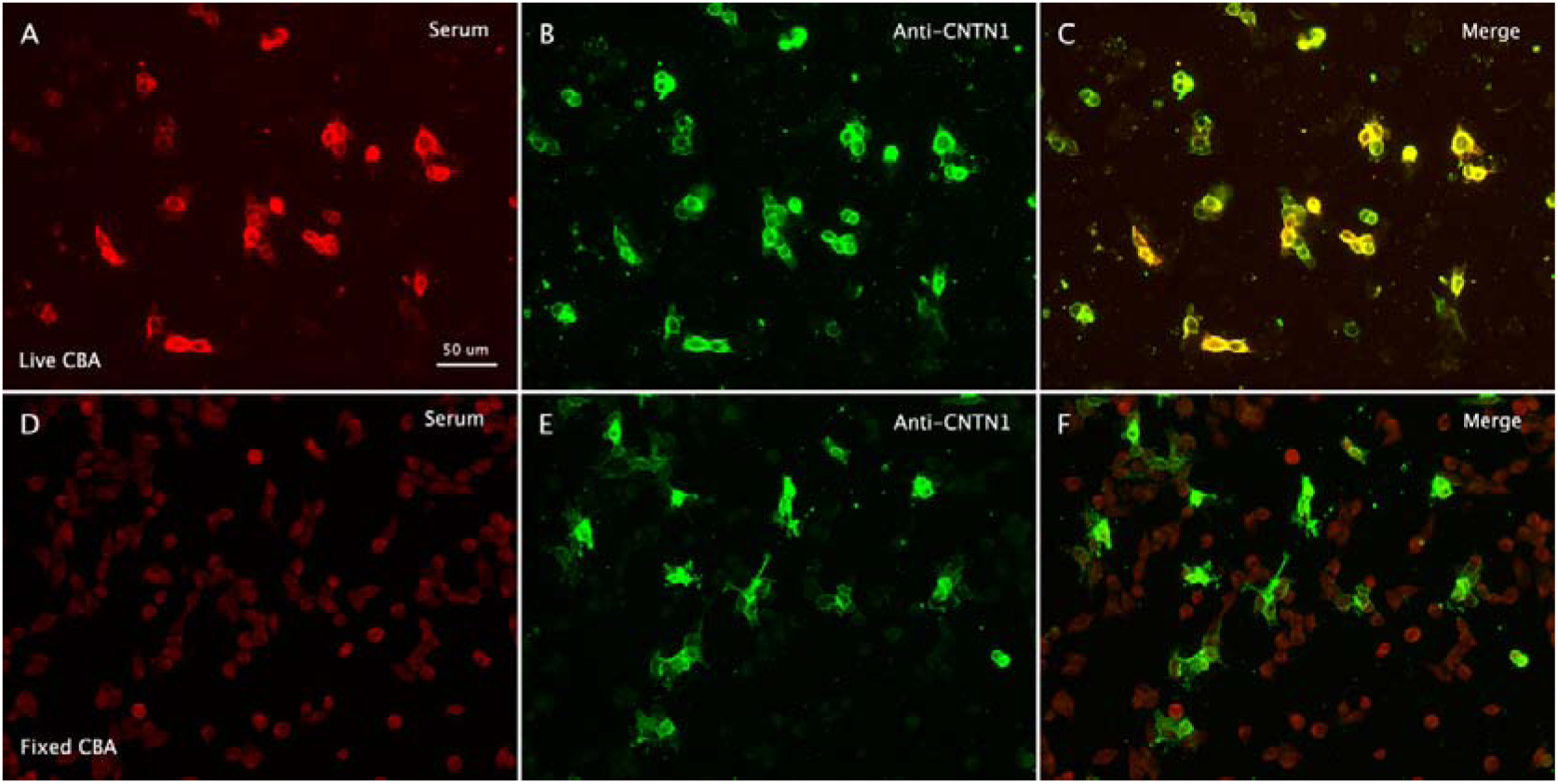
Live vs fixed CNTN1 CBA. Serum patient IgG staining (A,D) and commercial antibody against CNTN1 (B,E), in live (A,B,C) and fixed (D,E,F) CNTN1 CBA. One patient’s serum binds to CNTN1-trasnfected HEK293 cells when incubating in live cells (A), and not in fixed cells (D).

